# Nursing students’ attitudes, knowledge, and willingness to receive the COVID-19 vaccine: A cross-sectional study

**DOI:** 10.1101/2021.05.24.21257710

**Authors:** Ning Jiang, Baojian Wei, Hua Lin, Youjuan Wang, Shouxia Chai, Wei Liu

**Affiliations:** School of Nursing, Shandong First Medical University & Shandong Academy of Medical Sciences, Taian 271000, Shandong, China; School of Nursing, Hubei University of Medicine, Shiyan 442000, Hubei, China

**Author notes:** Corresponding author infromation: Wei Liu, RN, Professor, MD, School of Nursing, Shandong First Medical University & Shandong Academy of Medical Sciences, Taian 271000, Shandong, China. Ning Jiang, Baojian Wei and Hua Lin are co-first authors.

**Keywords:** COVID-19 vaccine, Attitudes, Knowledge, Willingness, Nursing students

## Abstract

**Aim:** To investigate nursing students’ konwledge, attitudes and willingness to receive the COVID-19 vaccine, and the influencing factors.

**Background:** Vaccination is one of the effective measures to prevent COVID-19, but the vaccination acceptance varies across countries and populations. As reserve nurses, nursing students have both the professionalism of medical personnel and the special characteristics of school students, their attitudes, knowledge, and willingness to receive the COVID-19 vaccine may greatly affect the vaccine acceptance of the population now and in the future. But little research has been done on vaccine acceptance among nursing students.

**Design:** A cross-sectional survey of nursing students was conducted via online questionnaires in March 2021.

**Methods:** Descriptive statistics, independent sample t tests/one-way ANOVA (normal distribution), Mann-Whitney U tests/Kruskal-Wallis H tests (skewness distribution) and multivariate linear regression were performed.

**Results:** The score rate of attitude, knowledge and vaccination willingness were 70.07%, 80.70% and 84.38% respectively. Attitude was significantly influenced by family economic conditions and whether a family member had been vaccinated. The main factors influencing knowledge were gender, grade and academic background. In terms of willingness, gender, academic background, visits to risk areas, whether family members were vaccinated, and whether they had side effects were significant influencing factors.

**Conclusions:** The vaccine acceptance of nursing students was fair. Greater focus needed to be placed on the males, those of younger age, with a science background, and having low grades, as well as on students whose family members had not received the COVID-19 vaccine or had side effects from the vaccine. Targeted intervention strategies were recommended to improve vaccination rates.

## 1 INTRODUCTION

The COVID-19 pandemic is a global public health crisis that has seriously impacted the international community. Vaccination is one of the most effective and low-cost measures to prevent COVID-19. Currently, nine COVID-19 vaccines have been approved for marketing worldwide, and as of March 15, 2021, more than 360 million doses of COVID-19 vaccines have been globally administered (WHO, 2021). Herd immunity is directly proportional to vaccination rate, and adequate population immunity can only be achieved if the vast majority of people are vaccinated. However, studies have shown that the willingness to be vaccinated varies across countries and populations after the introduction of the COVID-19 vaccine and is influenced by a number of factors. Kreps et al. (2020) conducted a survey of adults in the United States, with 69% of the participants found to be willing to accept the COVID-19 vaccine. They were more willing to receive the vaccine if the healthcare provider recommended it and if the perceived risk and severity of the disease were higher than those of vaccine side effects. Vaccines with high effectiveness, long protection periods, and low incidence of adverse reactions were easily accepted by the population (Kreps et al., 2020; Reiter, Pennell & Katz. 2020). Lin et al. (2020) conducted a survey of 3,541 residents in China on vaccination intention and willingness to pay and found that 83.3% of the residents were willing to be vaccinated and that willingness to be vaccinated was influenced by socioeconomic factors. Yuda and Katsuyama (2021) surveyed 1,100 Japanese residents and found that 65.7% of participants were willing to be vaccinated, with higher willingness to be vaccinated among the elderly, those in rural areas, and those with underlying diseases, but higher hesitation was noted among women. Among healthcare workers, the vaccine acceptance rate ranged from 23.4% in Taiwan to 95% in the Asia-Pacific region (Sallam, 2021; Szmyd et al., 2021; Kukreti et al., 2021; Shaw et al., 2021; Unroe et al., 2021). Side effects after vaccination (Szmyd et al., 2021), long-term protective efficacy and safety (Shaw et al., 2021) were the main concerns of healthcare workers. Depression (Szmyd et al., 2021), anxiety (Yurttas et al., 2021), and concerns about side effects (Unroe et al., 2021) were common reasons for reducing healthcare workers’ willingness to be vaccinated, while disease prevention (Kukreti et al., 2021), fear of transmitting disease to family members (Szmyd et al., 2021), clinical workplace (Shaw et al., 2021), older age, male, (Yurttas et al., 2021; Shaw et al., 2021; Unroe et al., 2021) white (Yurttas et al., 2021; Unroe et al., 2021), and prosociality (Sun & Operario, 2020) were the main reasons for promoting vaccination. In college students, Qiao et al. (2020) surveyed 1,062 South Carolina college students and revealed that perception and fear of the outbreak were positively associated with high vaccine acceptance, while higher exposure risk and negative attitudes toward the vaccine were associated with low vaccine acceptance. Graupensperger et al. (2021) conducted an online survey involving 647 college students and found that 91.64% of students were willing to receive the vaccine, indicating a strong willingness to receive the vaccine. However, participants thought that other young people were less likely to receive the vaccine and did not think vaccination was important. Sun et al. (2020) surveyed 1,912 Chinese university students, with 64.01% of the participants indicating that they were willing to participate in a COVID-19 vaccine trial. Low socioeconomic status, female, perceived risk of contracting the disease, and prosocial behavior were the main factors promoting willingness to be vaccinated, while hesitation to sign the informed consent form, time required to participate in the study, and perceived social stigma of COVID-19 were the primary factors which hindered willingness to be vaccinated. In Di Giuseppe et al.’s (2021) vaccination willingness survey involving a university population in southern Italy, 84.1% of participants were willing to be vaccinated, while males, unmarried/cohabiting, faculty, trust in vaccine safety, and clarity on possible side effects were important predictors of acceptance of the COVID-19 vaccine. China is conducting free vaccination for all, and the willingness of the population to receive vaccines is key to achieving reasonable vaccination coverage. As future medical personnel who will be working in hospitals, nursing college students have both the professionalism of medical personnel and the special characteristics of school students. As reserve nurses, if they have better knowledge of the COVID-19 vaccine, they can use their personal expertise to educate their relatives and friends in the neighborhood, the patients they will serve after working in the hospital, and the public regarding the vaccine. Furthermore, as students, if they have good vaccination attitudes and behaviors, they can set an example for other faculty students to improve the COVID-19 vaccination rate. Therefore, it is significant to understand nursing students’ perceptions, attitudes, vaccination intentions, and related influencing factors of COVID-19 vaccines, which can assist educational institutions in developing effective interventions to increase vaccination rate. Upon reviewing the existing literature, we found that only a few studies have been conducted on college students’ vaccination intentions, with nursing college students being rarely studied; thus, an investigation is imperative. The aim of this cross-sectional study was to investigate Chinese nursing students’ knowledge, attitudes, and vaccination intentions toward the COVID-19 vaccine and to examine potential influencing factors to provide evidence for developing intervention strategies and improving vaccination rates.

## 2 Methods

### 2.1 Design

A convenience sampling method was adopted to select two medical schools within mainland China. Then following the cluster sampling method, nursing college students were then selected as the study population. This study was a cross-sectional electronic survey. Data were collected through a web-link of an online questionnaire during mid-March 2021.

### 2.2 Participants and questionnaire

In this survey, a total of 15 basic information items and 3 questionnaire dimensions were covered, giving rise to 31 variables for statistical analysis. According to the Kendall sample estimation method for multivariate analysis, the sample required should be 10-20 times as high as the number of variables(Wang, 1990). In this regard, the minimum sample size of this survey was about 310-620. Participants met the following criteria: full-time learning; major in nursing; informed consent. A total of 1488 nursing students participated in this study.

### 2.3 Questionnaire

Based on the Guidelines for COVID-19 Vaccination (1st edition) issued by National Health Commission of the People’s Republic of China [China NHC] (2021). WHO’s ‘Vaccine Explained’ series features illustrated articles on vaccine development and distribution (2021), and guidelines of COVID-19 vaccine from the Chinese Center for Disease Control and Prevention (China CDC) (2021), the New York State Department of Health (2021), U.S. Food and Drug Administration [FDA] (2021) and the related literature (Qiao, Tam, & Li, 2020; Lin et al., 2020), we developed a questionnaire entitled “*Attitude,Knowledge and vaccination Willingness for* the *COVID-19 vaccine (AKW)”* which covered the following four parts:

1. Informed consent: including an introduction of the study, anonymity, confidentiality, guides for filling in the questionnaire and contact information.
2. Basic information (14 items): gender, age, grade, family status and vaccination experience, etc.
3. Attitude of the COVID-19 vaccine (11 items): influences of COVID-19, risk perception, vaccine acceptance, concern about the vaccine, etc. Each item was rated on a 5-point Likert scale and the total score ranged from 11-55, with a higher total score indicating more positive attitude.
4. Knowledge of the COVID-19 vaccine (9 items): (a)inoculation methods, suitable population, contraindications, adverse reactions, etc. The answer options included single choice and multiple choice. Each correct answer of single choice questions scored 5 and multiple choice questions scored 1 for each correct choice. The total score ranged from 1 to 46, with a higher total score indicating a better mastery of knowledge. (b) knowledge sources included mobile phone, TV, radio, network, newspaper, school/community pamphlet/bulletin board, relatives/friends introduce and others.
5. Vaccination Willingness (10 items): vaccine selection, vaccination form, willingness, reason, etc. Vaccination willingness was scored by 6-point Likert, the higher the score, the stronger the vaccination intention. The rest of the items were objective which described as percentages.

### 2.4 Data collection

We first called counselors (full-time faculty members who are responsible for managing students’ daily lives and studies in China) from the nursing schools of the two schools, trained them, and explained the purpose, content, and precautions of this study. The counselors then convened the students to explain this survey, and the students participated in the study after giving their informed consent. We uploaded the questionnaire to Wenjuanxing (https://www.wjx.cn), which is a commonly used online survey website in China. After uploading, a QR code poster was generated, and the researcher emailed the QR code poster to each counselor. They then uploaded the poster to student WeChat groups, and the students scanned the code to access Wenjuanxing website and fill out the questionnaire. IP address restriction technology was adopted to ensure users with the same IP address could only complete the questionnaire once. Data were downlouded from Wenjuanxing and questionnaires with up to 20% invalid entries were excluded.

### 2.5 Ethics aspects

The study was approved by the Human Subjects Ethics Sub-committee of Shandong First Medical University (registration number: R202105170156). All participants agreed to participate in the study. The questionnaire will be filled in anonymously, and the data will be kept strictly confidential for research purposes only.

### 2.6 Data analysis

The SPSS software (version 19.0; SPSS Inc.) was used for statistical analysis. Students’ demographic and information-sourcing characteristics were described as means with standard deviations (*SD*) and frequencies with percentages. The item scoring rate and the total scoring rate for AKW were calculated by dividing the actual score of an item or total items by the total item/items score and then multiplying by 100%. The data distribution was judged by histogram and Q-Q plot. Independent sample t tests/one-way ANOVA (normal distribution) and Mann-Whitney U tests/Kruskal-Wallis H tests (skewness distribution) were carried out to detect differences between participants in socio-demographic characteristics. Relationships among AKW were performed by Pearson or Spearman correlation analysis. Multiple linear regression was used to explore the influencing factors of the AKW. p < 0.05 was considered as statistically significant (two-tailed).

### 2.7 Validity and reliability

Four experts were invited to review the questionnaire, including two professors in public health, a community-based nursing specialist in charge of vaccination, and a nursing education specialist. All of the experts had more than 15 years of professional experience, and senior professional titles. The content validity index (I-CVI) for the questionnaire was 0.98. Prior to the survey, a convenience sampling method was used to select 20 nursing students who met the sampling criteria for a presurvey to clarify the acceptability of the questionnaire. The respondents found the questionnaire items clear and easy to understand. The Cronbach’s alpha coefficients for KAP scales was o.71.

## 3 RESULTS

A total of 1,512 copies of the questionnaire were returned, excluding those with incomplete baseline data and those with identical scores for all items, leading to a final number of 1,488 valid copies with an effectivity rate of 98.41%.

### 3.1 Characteristics of nursing students

The demographics of the study population that accounted for the majority were female (84.27%), between 21 and 22 years of age (23.99%), Han Chinese (98.52%), class of 2020 (42.14%), had a bachelor’s degree (82.93), from a science background in high school (60.15%), had parental family living in a new community (30.98%), League member (82.93%), were at a medium level of family economic status (78.49%), had a monthly consumption of 800-1,500 RMB (71.51%), had no history of travel to areas above medium risk in the past six months (99.26%), had no underlying diseases (97.98%), had no family members who received the COVID-19 vaccine (59.34%), and had a few family members who experienced side effects after receiving other vaccines (98.45%). (Table 1)

**Table 1.**
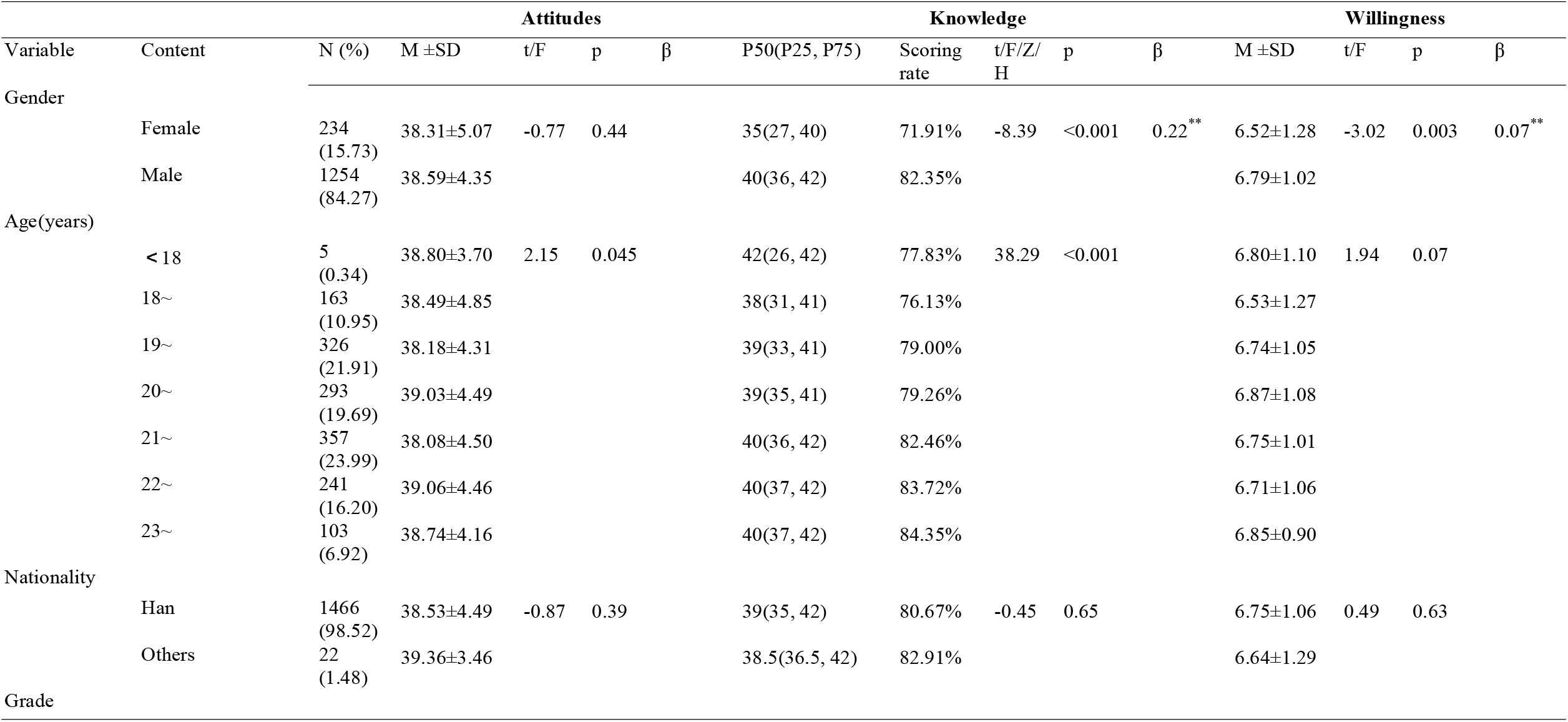
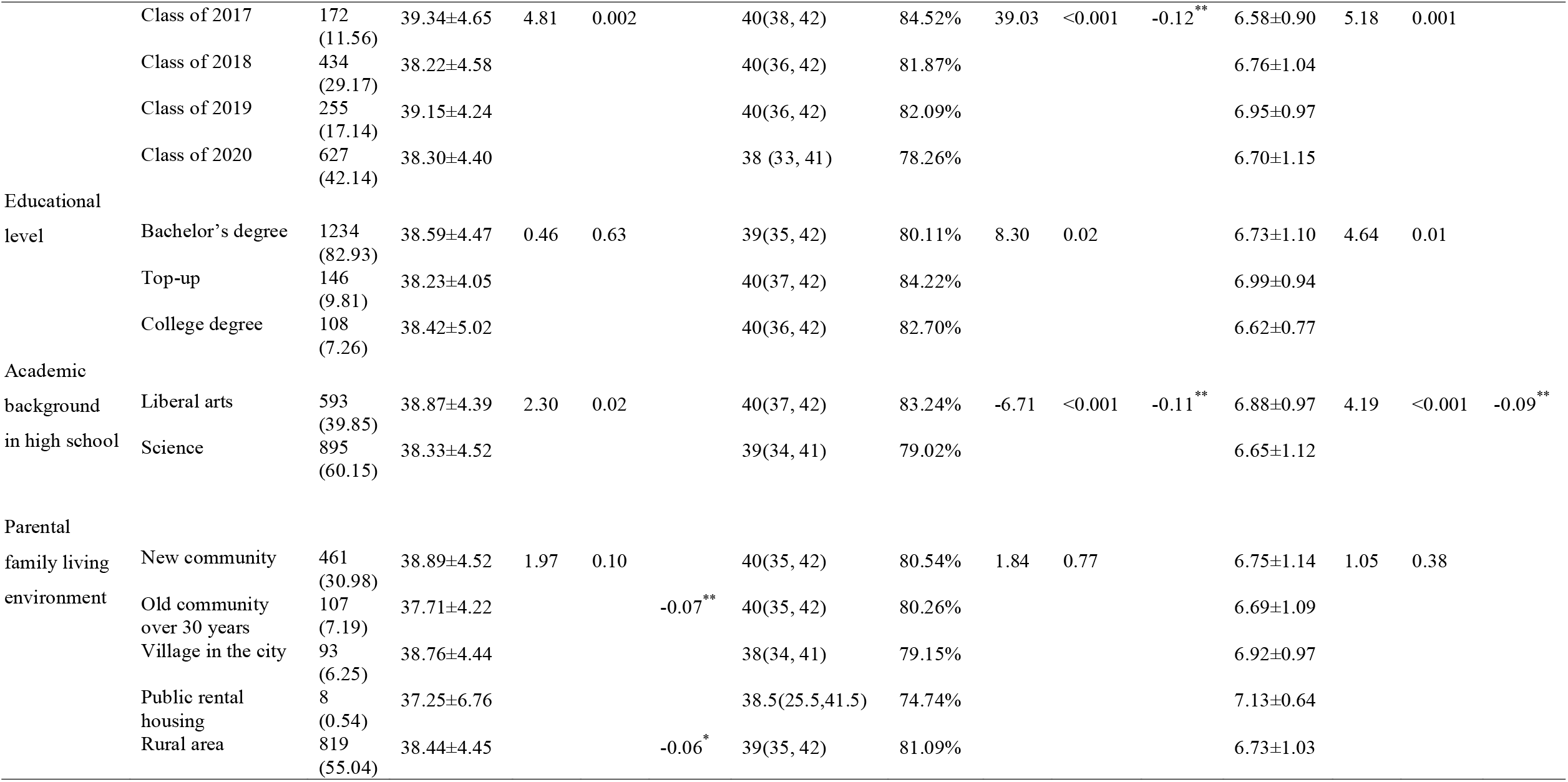
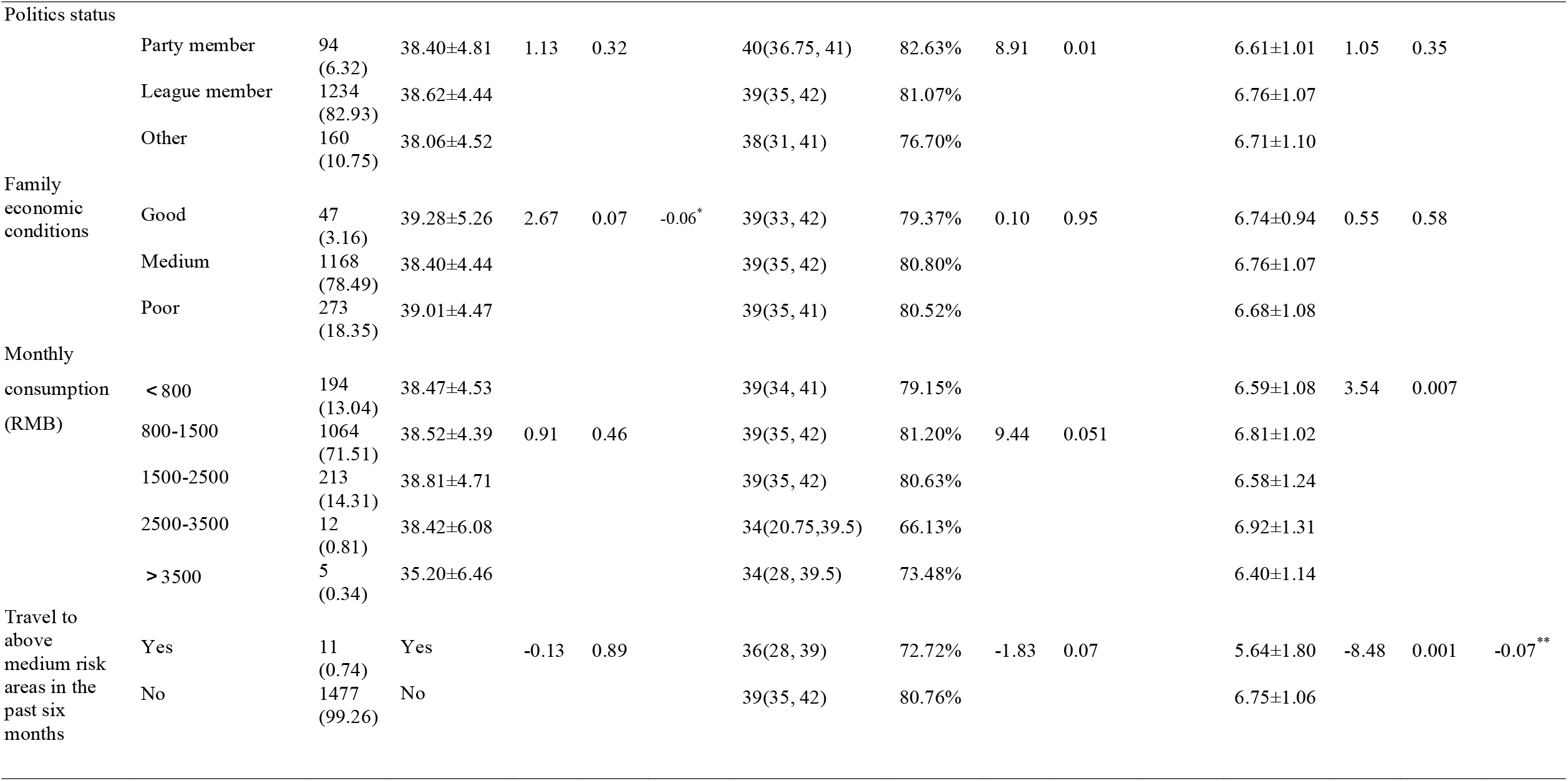
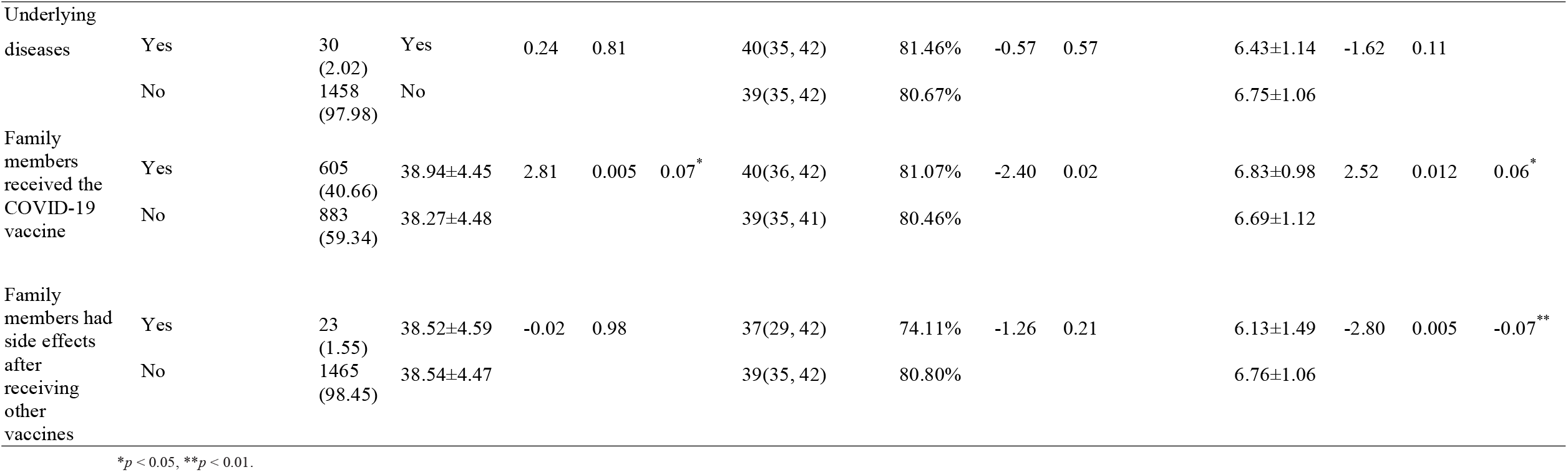
Demographics, univariate and multivariate analyses of factors associated with nursing students’ attitudes, knowledge, and willingness of COVID-19 vaccine (N = 1488)

### 3.2 Scoring

Attitude, willingness and total scal scores were normal distribution, and knowledge scores were skewed distribution. The mean score of the attitude dimension was 38.54 ± 4.48 and the score rate was 70.07%. In the attitude dimension, the highest score was for *Infection with COVID-19 has a high impact on the surrounding people or environment*. (88.80%), while the lowest score was for *Perceived high risk of infection with COVID-19*. (49.20%). The score rate in the knowledge dimension was 80.70%. In the knowledge dimension, *contraindications for the COVID-19 vaccine* (89.40%) had the highest score, while *correct method of vaccination* (63.75%) had the lowest score. The mean score of willingness to be vaccinated was 6.75 ± 1.07 with a score rate of 84.38%. The top three reasons for willingness to be vaccinated were as follows: responding to the national call (87.57%), believing in the vaccine (61.09%), and organizing group vaccinations at school (60.69%). The top three reasons for unwillingness to be vaccinated were as follows: worrying about the side effects of the vaccine (51.14%), the vaccine had just been introduced and its effect was unclear (42.41%), and observing the effect of vaccination on others (34.54%). In terms of vaccine selection, students were more willing to choose domestic vaccines (58.94%), and they hope that units or schools will organize collective vaccination (88.78%). The desired vaccine protection periods was more than 10 years (40.39%). Even with fees, 74.46% of students were still willing to be vaccinated, and the top three acceptable vaccine prices were within 50 RMB (40.99%), 5-100 RMB (36.29%), and 100-300 RMB (19.22%). (Table 2)

**Table 2.**
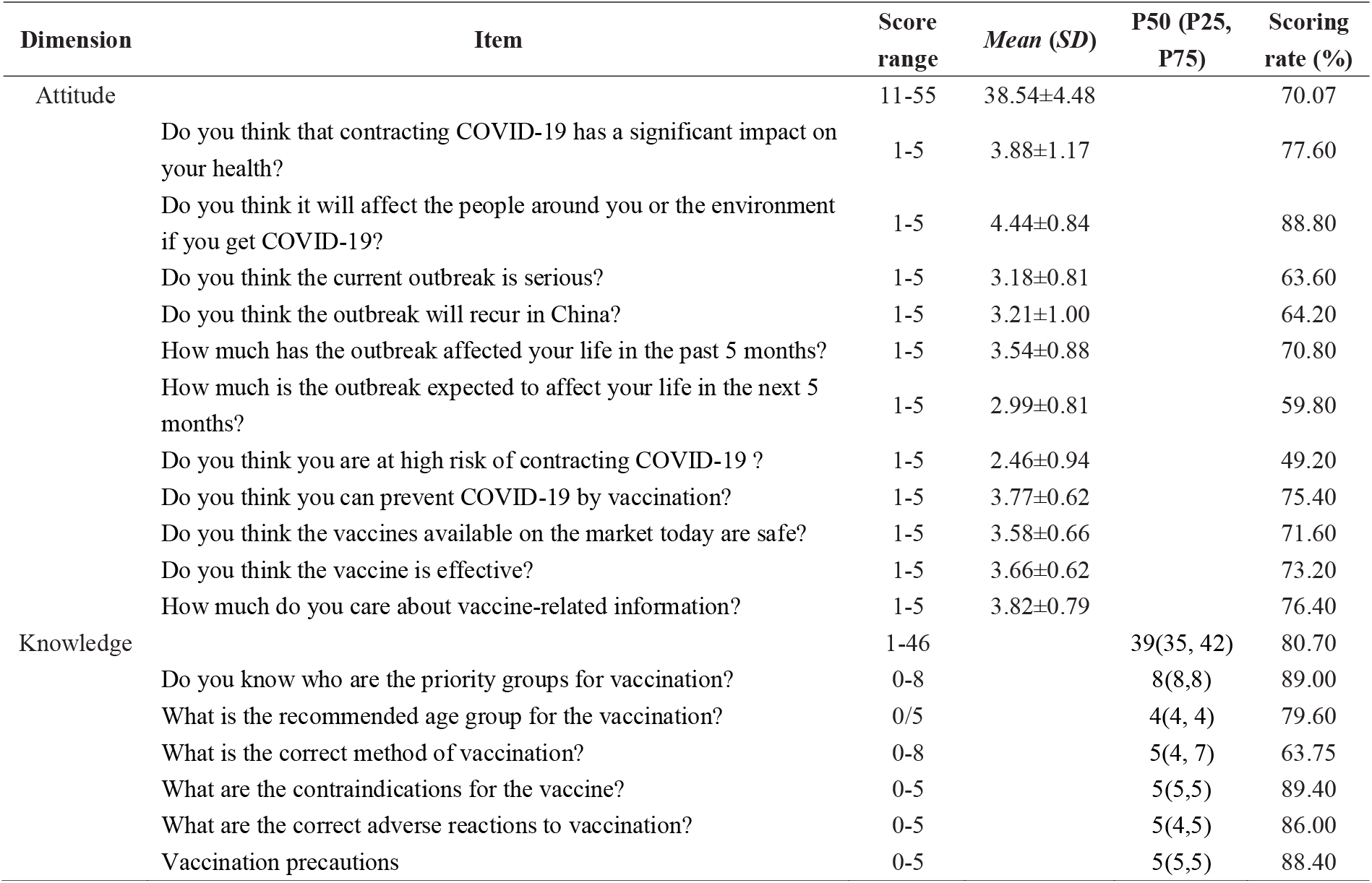
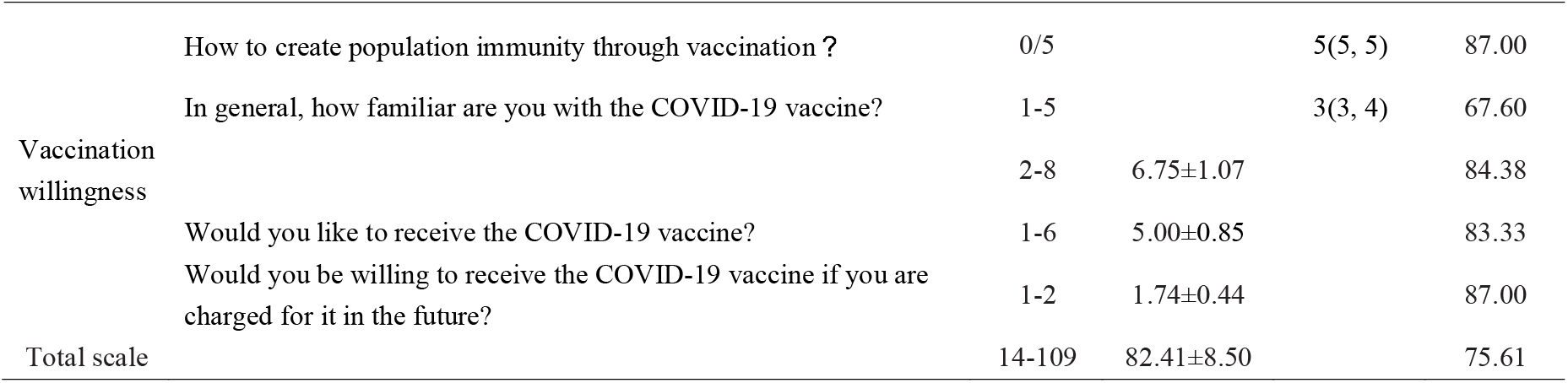
Scores of attitude, knowledge and willingness of COVID-19 vaccine (N = 1488)

**Figure 1.**
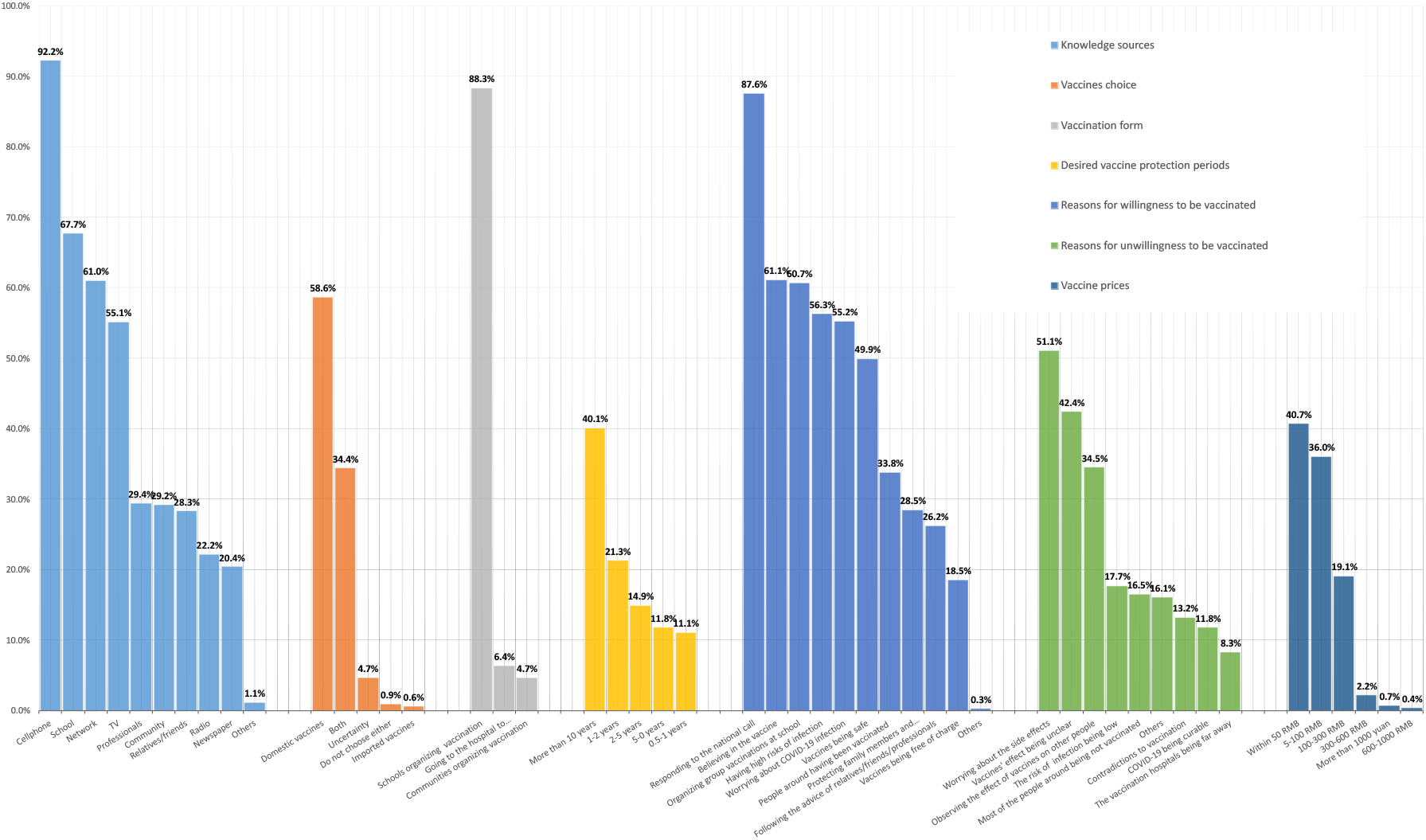
Knowledge sources, vaccines choice, vaccination form, desired vaccine protection periods, reasons for willingness and unwillingness to be vaccinated, vaccine prices

### 3.3 Influencing factors

The demographics characteristics of the subjects were used as grouping variables. From the statistical results, it is clear that when age, grade, academic background in high school, and family members who had received the COVID-19 vaccine were used as grouping variables, the differences in mean scores of attitudes were statistically significant, and that the attitudes toward vaccines were significantly higher among high-grade students than among low-grade students (juniors vs. sophomores and freshmen: 39.34 vs. 39.15 and 38.30, *p*=0.002). In terms of knowledge, the differences in scores were statistically significant in terms of sex, age, grade, educational level, academic background, politics status, and family members received the COVID-19 vaccine. The knowledge scores of female students were higher than those of male students (e.g. females vs. males: 82.35% vs. 71.91%, *p*<0.001), the knowledge mastery level increased significantly with age (e.g. 23∼, 22∼, 21∼ vs. 20∼, 19∼, 18∼: 84.35%, 83.72%, 82.46% vs. 79.26%, 79.00%, 76.13%, *p*<0.001), and the knowledge mastery level of top-up graduates was significantly higher than that of undergraduates and college graduates (e.g. top-up graduates vs. undergraduates and college graduates: 84.22% vs. 80.11% and 82.70%, *p*=0.02). With regard to vaccination willingness, there were significant differences in scores in terms of sex, age, grade, education level, parental living environment, family economic status, and monthly consumption. The willingness to be vaccinated was much higher among sophomores and juniors than among freshmen and seniors (e.g. sophomores and juniors vs. freshmen and seniors: 6.95 and 6.76 vs. 6.70 and 6.58, *p*=0.001), as well as among females versus males (e.g. females vs. males: 6.79 vs. 6.52, *p*<0.001).

Multiple linear regression analysis showed that family economic conditions and whether someone in the family had received the COVID-19 vaccine was a significant influence on attitude. The main factors influencing knowledge were gender, grade and academic background. Regarding the willingness to be vaccinated, gender, academic background, visits to areas above medium risk in the past six months, whether family members were vaccinated, and whether family members experienced side effects after receiving other vaccines were significant influencing factors. (Table 1)

## 4 DISCUSSION

This study, through a literature review, group discussion, and consultation with experts, constructed an attitudes, knowledge, and willingness (AKW) scale in terms of acceptance of the COVID-19 vaccine. The content validity of the scale was 0.98 and the Cronbach’s a coefficient was 0.71, indicating that the scale has some reliability. The AKW score in this study was 82.41±8.50, which was in the range of 14-109, indicating that nursing students had a positive perception of the COVID-19 vaccine and were willing to receive it.

### 4.1 Attitudes

low score items in the attitude dimension were *Do you think the current outbreak is serious?* (63.60%), *Do you think the outbreak will recur in China?* (64.20%), *How much is the outbreak expected to affect your life in the next 5 months?* (59.80%) and *Do you think you are at high risk of contracting COVID-19?* (49.20%). The related fact is that the pandemic is now better controlled in China, leading to students being less vigilant due to beliefs that the pandemic will not recur and that the chance of infection is small. This suggest that the acceptance of the vaccine was lower when the perceived severity and fear of COVID-19 were lower which is in agreement with the existing literature (Qiao, Tam & Li, 2020; Graffigna et al., 2020; Reiter, Pennell & Katz, 2020). Kwok et al. (2021) found that hospital nurses’ willingness to be vaccinated against COVID-19 declined after the situation regarding COVID-19 pandemic tended to improve. Therefore, it is recommended that schools should strengthen education regarding the COVID-19 vaccine and highlight the importance of vaccination among students, while accelerating the vaccination process with full respect for students’ vaccination willingness, to be able to increase vaccination rates. Although the risk of the pandemic is perceived as low, students also believe that once they infect COVID-19, it can have a significant impact on them personally and their surroundings. A positive attitude is an important factor in controlling the outbreak and receiving the COVID-19 vaccine (Yang et al., 2021). Family members receiving the COVID-19 vaccine had a positive influence on vaccination attitudes (Dror et al., 2020; Bell et al., 2020). Students in the present study had moderate approval of the vaccine’s effectiveness and safety, which was in line with existing studies (Kreps et al., 2020). Univariate analysis revealed that the attitude scores of the senior students were significantly higher than those of the other grades. It was suggested that as the senior students were in hospital internship, they would be exposed to various types of patients every day, and thus their risk of contracting COVID-19 was much higher than those of the other three grades. As a result, their attitudes toward the COVID-19 vaccine were more positive. Notably, students with liberal arts backgrounds scored higher on attitude. In China, nursing school mainly recruit science students, and liberal arts students, who account for a relatively small proportion in this survey, scored higher in attitude. The COVID-19 pandemic had a great psychological impact on liberal arts students than science students, who were more likely to experience anxiety (Odriozola-González et al., 2020), leading to greater awareness and recognition of the vaccine (Reiter, Pennell & Katz et al., 2020).

### 4.2 Knowledge

The scoring rate of knowledge dimension was 80.70%. Item analysis showed that out of the 7 knowledge items, 5 items (contraindication of vaccination, key vaccinated groups, precautions after vaccination, herd immunity, adverse reactions) scored at a rate of more than 80%, indicating that most students had a good grasp of the knowledge of COVID-19 vaccine. However, the relatively low scoring items of *the correct way to vaccinate* (63.75%) and *the recommended age group for the vaccination* (79.60%) suggested that these were weak points that needed to be strengthened. In the *correct method of vaccination* item, adenovirus vaccines and recombinant vaccine inoculation methods served the lowest. Only 40.39% of the students could clearly point out that nucleic acid testing was not necessary before vaccination, while most of the students believed that nucleic acid testing was necessary. Fear of the discomfort of nucleic acid testing might influence students’ willingness to vaccinate (Frazee et al., 2018). 69.83% of the students believed that the age group suitable for vaccination was 18-60 years old. However, according to the latest guidelines issued by the National Health Commission of China [China NHC] (2020), the recommended age for vaccination has been revised to be above 18 years old, which suggested that the knowledge students have mastered should be updated in time. In terms of vaccine information sources, mobile phones, school propaganda, computers and televisions ranked the top. Mobile phones and computers were the daily necessities for college students, which were the main sources of information. But interestingly, school propaganda performed better than computers in this study, which was not consistent with previous research (Olaimat et al., 2020). College students are an important group for epidemic prevention. China is now vigorously carrying out the policy of universal immunization, which has attracted great attention from government departments, school management and teachers. Schools often hold lectures on COVID-19, emphasizing the importance of COVID-19, and integrating COVID-19 related knowledge into students’ daily lives, which improved students’ knowledge of COVID-19 vaccine. Grade, gender and high school background were significant predictors of knowledge. Higher age and senior students were in a dominant position in acquiring and absorbing knowledge (Olaimat et al., 2020), so their knowledge scores were significantly higher than those of lower age and junior students. Girls can master better knowledge than boys. This gender difference may have something to do with nursing profession (Albaqawi et al., 2020), or with gender itself (Zhong et al., 2020). Therefore, nursing education must ensure that students of all grades and genders have equal access to student-centered COVID-19 resources to prevent knowledge biases among students.

### 4.3 Vaccination willingness

Among the 1,488 students interviewed, the scoring rate of vaccination willingness was 5.00±0.85 (83.33%), which was in the middle-to-upper level compared with existing studies (Qiao, Tam & Li, 2020; Sun, Lin & Operario, 2020; Graupensperger, Abdallah & Lee, 2021; Di Giuseppe et al., 2021), indicating that nursing students were more willing to be vaccinated. The analysis of the reasons for willingness/unwillingness to be vaccinated showed that nursing students could have a certain degree of understanding of the necessity, effectiveness, and safety of the COVID-19 vaccine based on their professional foundation. Moreover, their schools are preparing to organize group vaccinations, so that their vaccination willingness is high. What cannot be ignored is that students still have hesitations about safety and effectiveness. If these hesitations about vaccination are not addressed before vaccination, it may reduce the actual number of vaccinated students and cause psychological pressure as well as unnecessary distress after vaccination (Lucia, Kelekar & Afonso, 2020; Qiao, Tam & Li, 2020). According to the results of the regression analysis, sex, academic background, visits to areas above medium risk in the last six months, whether family members were vaccinated, and whether they had side effects after vaccination were significant influencing factors for the vaccination willingness. In this study, the vaccination willingness of female students was significantly higher than that of male students, which is inconsistent with part of existing studies (Freeman et al., 2020; Unroe et al., 2021). The reason may be that nursing is traditionally a female-dominated profession, and male students who are a minority group in nursing schools sometimes feeling ashamed or isolated and are reluctant to participate in group activities (Power et al., 2018), which may lead to a decrease in vaccination willingness (Christensen, Welch & Bar, 2018). Therefore, it is worthwhile for nursing educators to think about putting themselves in the shoes of different types of students and paying attention to the small but important group of male students during vaccination. In this study, the vaccination rate of family members of the surveyed students was 40.66% (605/1488), and the incidence of adverse reactions was 6.12% (37/605), which was higher than the median incidence of adverse reactions in existing studies (0.1-15.8%) (Baden et al., 2021; China NHC, 2021). Therefore, students may have negative cognition in vaccination. In view of this, schools should strengthen education on adverse reactions of COVID-19 vaccine, not only from theoretical knowledge, but also from psychological, physiological, so that students can have an objective understanding of the side effects of vaccine, so as to eliminate fear and enhance the willingness to vaccinate. Concerning vaccine selection, most students are willing to receive the vaccine even with a fee, with the acceptable price being within 100 RMB. Most of the students in this study spend 800-1,500 RMB per month. Thus, a vaccine price within 100 RMB is affordable for students. This also provides a reference for policy makers when setting vaccine prices, with recommendations that the vaccine prices be reasonable by basing it on the premise of ensuring medical equity.

### 4.4 Limitations

There are some limitations to this study. First, to ensure the timeliness of the survey, we conducted an initial validation of the survey instrument through expert review. Although the CVI coefficient and Cronbach’s alpha coefficient are acceptable, further standard validation measures are needed. Second, the sample of this study was mainly taken from two undergraduate schools having mostly nursing undergraduates and was based on the enrollment level of the schools as well as the students’ willingness to fill out the survey; thus, the results need to be interpreted with caution. Third, this study was conducted three months after the vaccine was released, which may only reflect initial willingness to be vaccinated. Further research is still needed to analyze mid- and late-stage willingness to be vaccinated as well as actual vaccination behavior.

## 5 CONCLUSION

The study initially explored nursing students’ attitudes, knowledge, and vaccination willingness as well as related factors. The nursing students were not only representative of medical professionals, but also representative of college students. It provided a reference for policy makers to understand the vaccination willingness of medical workers and young people which could optimize the management strategy of neo-crown vaccination. And for nursing educators to enhance follow-up education and increase vaccination rates, with a focus on conducting interventions for male students, lower age groups, those with science backgrounds and lower grades, students whose family members have not received the COVID-19 vaccine, as well as those whose family members have experienced vaccine side effects. Due to the limitations posed by the study’s sample, further expansion of the study population to include post-secondary and specialist nursing students is needed to support our findings. In addition, further studies related to students’ anonymous vaccination intentions and actual vaccination behaviors are needed in order to evaluate the practical significance of this survey.

## Data Availability

All data referred to in the manuscript could be obtained by http://gofile.me/53Bpp/sywG4e20H

## ACKNOWLEDGMENTS

The authors would like to acknowledge all nursing students and their counselors in this study for their contributions.

